# Universal screening for SARS-CoV-2 infection among pregnant women at Elmhurst Hospital Center, Queens, New York

**DOI:** 10.1101/2020.08.12.20171694

**Authors:** Sheela Maru, Uday Patil, Rachel Carroll-Bennett, Aaron Baum, Tracy Bohn-Hemmerdinger, Andrew Ditchik, Michael L Scanlon, Parvathy Krishnan, Kelly Bogaert, Carson Woodbury, Duncan Maru, Lawrence Noble, Randi Wasserman, Barry Brown, Rachel Vreeman, Joseph Masci

**Affiliations:** Department of Health System Design and Global Health and the Arnhold Institute for Global Health, Icahn School of Medicine at Mount Sinai, New York City, NY, USA; Department of Obstetrics, Gynecology and Reproductive Science at the Icahn School of Medicine at Mount Sinai, New York City, NY, USA; New York City Health + Hospitals/Elmhurst, New York City, NY, USA; Department of Pediatrics, Icahn School of Medicine at Mount Sinai, New York City, NY, USA; Department of Medicine, Icahn School of Medicine at Mount Sinai, New York City, NY, USA

## Abstract

**Background:** Universal screening for SARS-CoV-2 infection on Labor and Delivery (L&D) units is a critical strategy to manage patient and health worker safety, especially in a vulnerable high-prevalence community. We describe the results of a SARS-CoV-2 universal screening program at the L&D Unit at Elmhurst Hospital in Queens, NY, a 545-bed public hospital serving a diverse, largely immigrant and low-income patient population and an epicenter of the global pandemic.

**Methods and findings:** We conducted a retrospective cross-sectional study. All pregnant women admitted to the L&D Unit of Elmhurst Hospital from March 29, 2020 to April 22, 2020 were included for analysis. The primary outcomes of the study were: (1) SARS-CoV-2 positivity among universally screened pregnant women, stratified by demographic characteristics, maternal comorbidities, and delivery outcomes; and (2) Symptomatic or asymptomatic presentation at the time of testing among SARS-CoV-2 positive women.

A total of 126 obstetric patients were screened for SARS-CoV-2 between March 29 and April 22. Of these, 37% were positive. Of the women who tested positive, 72% were asymptomatic at the time of testing. Patients who tested positive for SARS-CoV-2 were more likely to be of Hispanic ethnicity (unadjusted difference 24.4 percentage points, CI 7.9, 41.0) and report their primary language as Spanish (unadjusted difference 32.9 percentage points, CI 15.8, 49.9) than patients who tested negative.

**Conclusions:** In this retrospective cross-sectional study of data from a universal SARS-Cov-2 screening program implemented in the L&D unit of a safety-net hospital in Queens, New York, we found over one-third of pregnant women testing positive, the majority of those asymptomatic. The rationale for universal screening at the L&D Unit at Elmhurst Hospital was to ensure safety of patients and staff during an acute surge in SARS-Cov-2 infections through appropriate identification and isolation of pregnant women with positive test results. Women were roomed by their SARS-CoV-2 status given increasing space limitations. In addition, postpartum counseling was tailored to infection status. We quickly established discharge counseling and follow-up protocols tailored to their specific social needs. The experience at Elmhurst Hospital is instructive for other L&D units serving vulnerable populations and for pandemic preparedness.

## Introduction

New York City (NYC) has been a global epicenter of the SARS-CoV-2 outbreak, accounting for 17% of confirmed cases in the United States, as of April 25, 2020 [1]. In the initial response, SARS-CoV-2 testing in NYC focused on symptomatic individuals at testing centers or emergency departments and hospitalized patients [2]. Asymptomatic infected individuals may contribute substantially to transmission, including in hospital settings [3-9]. The fraction of SARS-CoV-2 infections in NYC that are asymptomatic – a key parameter for epidemiological models that forecast rates of infection, hospitalization, and death – is unknown. Moreover, much of the available data is not stratified by socio-demographic characteristics, which is critical to understand health disparities [10].

Labor and delivery (L&D) wards were some of the first places where universal screening for SARS-CoV-2 was instituted in the US. On March 29, 2020, the L&D Unit at Elmhurst Hospital in Queens, NYC instituted universal screening in order to manage care and isolation of admitted patients. The primary objective of this study was to describe the prevalence of SARS-CoV-2 infection, stratified by socio-demographic data, and symptom presentation among universally screened pregnant women. We explain the rationale, implications, and significance of screening in this public hospital caring for a severely affected and vulnerable population.

## Methods

### Study design and setting

We retrospectively reviewed medical charts of all pregnant women who were universally screened for SARS-CoV-2 on admission to the L&D Unit at Elmhurst Hospital. Elmhurst Hospital is a 545-bed public hospital that serves a diverse, largely immigrant and low-income patient population. The hospital serves about 134,000 patients per year and performs 2,200 deliveries annually [11]. The hospital has been hard hit by the pandemic, caring for 2,920 known COVID-19 cases as of May 4, 2020 [12].

### Participants and data collection

We included all pregnant women admitted to the L&D Unit at Elmhurst Hospital from March 29, 2020 to April 22, 2020. This time period was selected as it comprised the ‘surge’ of cases in NYC. Study size was limited by the time period. Demographic and clinical data were extracted from the electronic medical record (EPIC) both using a data extraction tool (SlicerDicer) and manually, then de-identified manually and assigned a random study number. SARS-CoV-2 testing was performed using nasopharyngeal swabs and the results were from BioReference Laboratories and the Elmhurst Hospital Laboratory (Cepheid Rapid PCR). Primary outcomes were SARS-CoV-2 test result (positive/negative/invalid) and the presence of SARS-CoV-2 symptoms on admission. Demographic data included age, ethnicity, primary language, marital status (‘not married’ includes single and divorced statuses), health insurance status (public, private and uninsured), and zip code. Race was not included due to the poor quality of the data in the medical record. Clinical data included SARS-CoV-2 test result, SARS-CoV-2 symptoms, gestational age at delivery (divided into term and preterm), mode of delivery (vaginal delivery includes assisted vaginal deliveries), comorbidities, and length of stay. The comorbidities we assessed were as follows: hypertensive disorders (chronic hypertension, gestational hypertension, preeclampsia, superimposed preeclampsia), pre-pregnancy maternal obesity, asthma or pulmonary disease, diabetes (pre-gestational and gestational), infectious disease (HIV, hepatitis B), depression or anxiety, and other significant maternal disease including heart, kidney, or thyroid disease.

### Statistical analysis

We describe SARS-CoV-2 test positivity and the symptom status at admission among positive patients. Test positivity is stratified by sociodemographic characteristics, maternal comorbidities, and delivery outcomes. Demographic characteristics, maternal comorbidities, and delivery outcomes of patients who tested positive versus negative for SARS-CoV-2 were compared using 2-sided unpaired t-tests, with unadjusted differences and 95% confidence intervals (CI) reported.

### Ethical approval

This study was approved by the Institutional Review Board at the Icahn School of Medicine at Mount Sinai, IRB-20-03424, and approved by NYC Health + Hospitals.

## Results

One hundred percent of admissions to the L&D Unit during the study period were tested. Two women had invalid test results and were excluded from analysis. Among the remaining 124 women, the mean age was 30.2 years, 78 (62.9%) were Hispanic, 26 (20.1%) were Asian, and 112 (90.3%) had public insurance. 78.3% of patients who tested positive for SARS-CoV-2 identified their ethnicity as Hispanic compared to 53.9% of patients who tested negative, a significant difference of 24.4 percentage points (95% CI: 7.9, 41.0). Similarly, 73.9% of patients who tested positive for SARS-CoV-2 self-reported their primary language as Spanish compared to 41.0% of patients who tested negative, a significant difference of 32.9 percentage points (95% CI: 15.8, 49.9). There was not a statistically significant difference in other demographic characteristics or rates of maternal comorbidities between patients who tested positive versus negative for SARS-CoV-2 (Table 1).

**Table 1.** SARS-CoV-2 Test Results among 124 Obstetrical Patients Admitted to Labor and Delivery, by Demographic Characteristics.

| Variable | SARS-CoV-2<br>Positive<br>No. (%) | SARS-CoV-2<br>Negative<br>No. (%) | Unadjusted Difference<br>(CI) |
| --- | --- | --- | --- |
| **Total Number** | 46 | 78 | -- |
| **Age (years)** |  |  |  |
| < median (30 years) | 20 (43.5%) | 43 (55.1%) | -11.6 (-30.1, 6.7) |
| > median (30 years) | 26 (56.5%) | 35 (44.9%) |  |
| **Ethnicity** |  |  |  |
| Hispanic | 36 (78.3%) | 42 (53.9%) | 24.4 (7.9, 41.0) |
| Not Hispanic | 10 (21.7%) | 36 (46.2%) |  |
| **Primary Language** |  |  |  |
| Spanish | 34 (73.9%) | 32 (41.0%) | 32.9 (15.8, 49.9) |
| Other | 12 (26.1%) | 46 (59.0%) |  |
| **Marital status** |  |  |  |
| Married | 19 (41.3%) | 36 (46.2%) | -4.8 (-23.2, 13.5) |
| Not Married | 27 (58.7%) | 42 (53.9%) |  |
| **Insurance status** |  |  |  |
| Public | 43 (93.5%) | 69 (88.5%) | 5.0 (-5.2, 15.3) |
| **Other** | 3 (6.5%) | 9 (11.5%) |  |
| **Comorbidities** |  |  |  |
| **≥ 1** | 26 (56.5%) | 35 (44.9%) | 11.6 (-6.8, 30.1) |
| **None** | 20 (43.5%) | 43 (55.1%) |  |
| **Current or Prior Smoker** |  |  |  |
| **Yes** | 2 (4.4%) | 5 (6.4%) | -2.1 (-10.2, 6.1) |
| **No** | 44 (95.7%) | 73 (93.6%) |  |

Forty-six of 124 (37.1%) screened pregnant women had a positive SARS-CoV-2 test. Of those, 33 (71.7%) were asymptomatic and 13 (28.3%) were symptomatic (Fig 1). Of those women who were symptomatic, the median number of symptoms on admission was 2, with the most common symptoms fever and cough. Other symptoms that were reported by patients in this cohort included body aches, sore throat, shortness of breath or difficulty breathing, headache, and runny nose. The presence of symptoms did not appear to be related to the mode of delivery or associated with preterm delivery. Of note, during our study period there were no maternal deaths.

**Fig 1.**
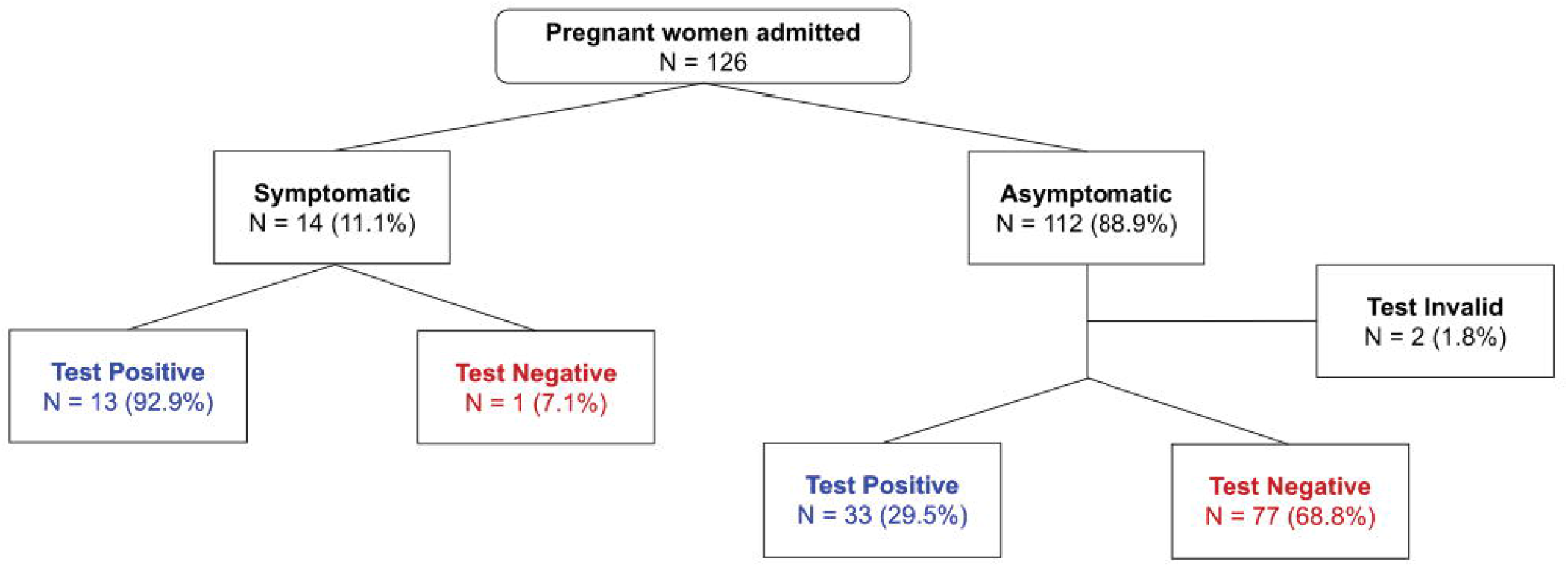
SARS-CoV-2 Symptoms on Admission and Test Results among 126 Obstetrical Patients Presenting for Delivery.

Among 38 SARS-CoV-2 positive women who delivered by the time of data collection, 35 (92.1%) had a term delivery and 25 (65.8%) had a vaginal delivery. This was similar to 74 SARS-CoV-2 negative women who delivered by the time of data collection, among whom 78 (87.8%) had a term delivery and 49 (66.2%) had a vaginal delivery. A significantly greater percentage of women who tested positive for SARS-CoV-2 had a length of stay greater than or equal to 48 hours compared to women who tested negative (45.7% vs 25.6%, unadjusted difference of 20.0 percentage points, 95% CI: 2.3, 37.7). (Table 2).

**Table 2.** SARS-CoV-2 Test Results among 124 Obstetrical Patients Admitted to Labor and Delivery, by Delivery Outcome.

| Variable | SARS-CoV-2<br>Positive<br>No. (%) | SARS-CoV-2<br>Negative<br>No. (%) | Unadjusted<br>Difference<br>(CI) |
| --- | --- | --- | --- |
| **Total Number** | 46 | 78 | -- |
| **Gestational Age <sup>a,b</sup>** |  |  |  |
| **Term** | 35 (92.1%) | 65 (87.8%) | 4.3 (-7.3, 15.9) |
| **Preterm** | 3 (7.9%) | 9 (12.2%) |  |
| **Mode of Delivery <sup>b</sup>** |  |  |  |
| **Vaginal** | 25 (65.8%) | 49 (66.2%) | 0.4 (-18.5, 19.3) |
| **Cesarean** | 13 (34.2%) | 25 (33.8%) |  |
| **Length of Stay** |  |  |  |
| **≥48 hours** | 21 (45.7%) | 20 (25.6%) | 20.0 (2.3, 37.7) |
| **<48 hours** | 25 (54.4%) | 58 (74.4%) |  |

<sup>a</sup> At time of delivery.
<sup>b</sup> Excludes 12 patients who were still pregnant at the time of data collection.

Seventy-eight of the screened women resided in one of 4 zip-codes, with the remaining residing across 29 distinct zip-codes. The NYC Department of Health and Mental Hygiene reported that, as of April 22, 2020, these 4 zip-codes had a SARS-CoV-2 test-positive rate of 59% to 79% for the general population [9]. In our data, women residing in these top four zip-codes had a test-positive rate of 30.4%.

## Discussion

Our study represents one of the few published studies of universal screening for SARS-CoV-2. We found that over one third of pregnant women tested positive for SARS-CoV-2 at the L&D Unit at Elmhurst. Selection bias may have been present as pregnant women with SARS-CoV-2 infection or symptoms may have been more likely to present for admission. However, this is significantly higher than results published from universal screening conducted at L&D Units in other NYC hospitals [8, 13-14]. A study among 215 pregnant women at New York-Presbyterian Allen Hospital and Columbia University Irving Medical Center found that 15% tested positive for SARS-CoV-2, and another among 161 pregnant women at NYU Winthrop Hospital found that 20% tested positive for SARS-CoV-2.

The higher rate of infection at Elmhurst Hospital is likely secondary to the demographics of the surrounding population. Over 200,000 Queens residents live in crowded homes, and 70,000 in severely crowded homes [15]. Recent data from NYC show a lower rate of infection for non-hospitalized Hispanic patients compared with Black or White patients; however, these data on race and ethnicity that come from laboratory reports were noted to be only 40% complete. Our universal screening data show significantly higher rates of infection among Hispanics compared to non-Hispanics, which is more consistent with data among all hospitalized patients in NYC [16]. Gaps in data are likely obscuring disparities.

The cross-sectional association we observed between test-positivity and Hispanic ethnicity and Spanish as a primary language may be confounded by patients’ socioeconomic status. At Elmhurst Hospital we serve a largely immigrant (foreign-born), resource-poor population. The primary language of Spanish likely indicates the patient was not born in the USA or is a first-degree descendant of Latin American immigrants (although there may be exceptions). Both of these characteristics are common in our patient population. Additionally, Spanish as a preferred language may indicate barriers to seeking or utilizing healthcare, which may affect co-morbidities and infection rates.

When looking at mode of delivery and gestational age at delivery, no significant difference in either of these was noted between the SARS-CoV-2 positive and negative mothers. SARS-CoV-2 status was not an indication for cesarean section or for preterm induction of labor.

The increased length of stay in the symptomatic patients is likely secondary to the severity of illness being greater in symptomatic patients. In order to warrant admission for SARS-Cov-2 symptoms typically maternal fever had to be accompanied by either maternal or fetal tachycardia, shortness of breath or decreased oxygen saturation on room air, or concerning findings on chest X-ray for possible pneumonia. Patients who did not meet these criteria, and had normal vital signs apart from fever, and reassuring fetal testing were typically assessed in our triage area and discharged home rather than admitted. These patients who were not admitted would not have been included in this cohort. The criteria for discharge among symptomatic patients included normalization of vital signs, specifically remaining afebrile and having oxygen saturation of over 95% consistently on room air both at rest and with ambulation.

Our first Obstetric SARS-CoV-2 case was March 15. Early in the pandemic, when we tested based on symptoms and history alone, several obstetric patients were admitted for birth asymptomatic and were later readmitted with severe COVID-19 infection. We were concerned about the likely spread to staff and other patients within our unit from pre-symptomatic and asymptomatic women. At the same time, our hospital was quickly becoming the epicenter of cases in the US, and all available physical space was required to care for COVID-19 patients. A large part of the postpartum unit was needed for non-obstetric COVID-19 care, and we suddenly had limited capacity to isolate positive postpartum women and those with unknown status. Upon this structural change, we began universal screening of pregnant women with planned or unplanned admission. One limitation of our study was the use of two different test types for universal screening. We were initially using BioReference Laboratory tests which returned in several days, and as rapid tests became available we switched to the Cepheid Rapid PCR. These tests likely had different sensitivities and specificities (though we were not provided with this information) and may have affected our screening results.

SARS-CoV-2 screening fundamentally shifted the way we roomed, counseled, and followed women (Fig 2). Cohorting women by SARS-CoV-2 infection status allowed us to most safely use the restricted space. Several walls were built to further partition the space in the triage, labor, and postpartum areas in order to ensure separation of SARS-CoV-2 positive women and those of unknown status from women known to be negative. In the early days of the pandemic, SARS-CoV-2 positive mothers were roomed separately from their infants, however, this procedure was reconsidered as the volume of known SARS-CoV-2 positive mothers increased with universal screening. Given limited physical space and staff to isolate all infants and considering the importance of simulating the home-environment during the postpartum hospitalization to promote safe infant care and breastfeeding, we proceeded with shared decision making with SARS-CoV-2 positive mothers around rooming-in with their infants. Severe overcrowding of housing is a challenge where most of our mothers live, making isolation of a baby and mother in the home-setting infeasible for most. We educated women on appropriate PPE use, breastfeeding, and social distancing measures that families could utilize upon discharge home. We postponed postpartum maternal vaccinations for SARS-CoV-2 positive women to eliminate any confusion between vaccine-related fever and COVID-19 symptoms (those who were SARS-CoV-2 negative received routine postpartum vaccination). Additionally, upon discharge we provided infected and PUI (Person Under Investigation) mothers with surgical facemasks and single-use Tempa-DOT thermometers, as the pharmacies in the surrounding zip-codes had none available. Our clinicians actively followed our infected and PUI mothers via phone after discharge, screening for symptoms and access to thermometers and acetaminophen.

**Fig 2.**
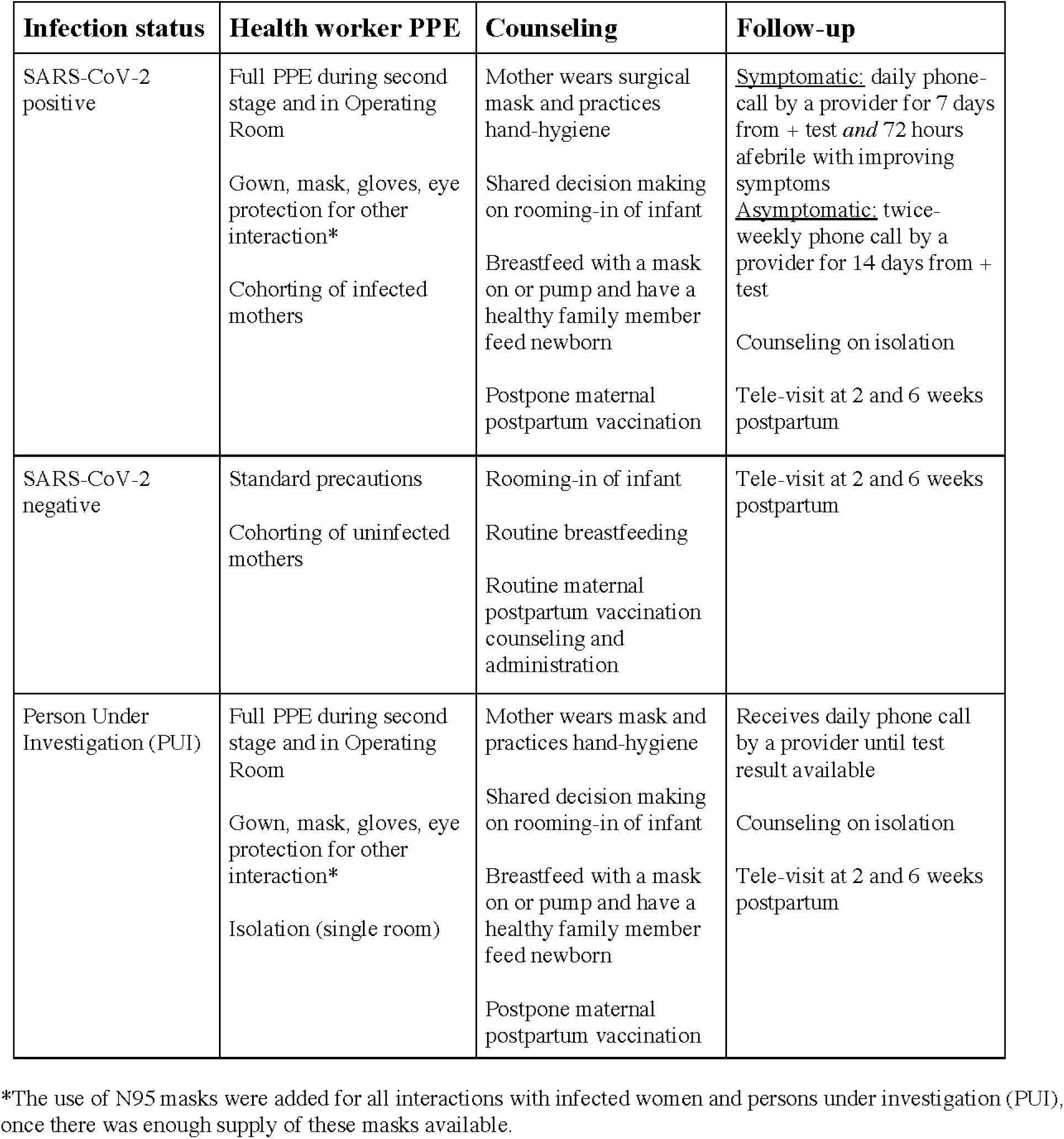
Strategies Employed by the Labor and Delivery Unit at Elmhurst Hospital during the Initial Phase of the SARS-CoV-2 Outbreak.

We found high rates of asymptomatic SARS-CoV-2 infection among pregnant women; these screening data and our experience at a large public hospital at the epicenter of the nation’s pandemic are instructive. In future epidemics, it may be prudent to look at L&D screening numbers early on, as pregnant women continue to seek essential care despite social distancing measures and also represent the general young and healthy community population. Additionally, this experience illustrates a nimble response to an acute shortage of space and PPE for hard-hit vulnerable communities.

## Data Availability

We are currently obtaining IRB review of the de-identified dataset prior to making it publicly available.
Please contact for more information.

## Acknowledgements

We would like to acknowledge the staff of Obstetrics and Gynecology and Pediatrics at Elmhurst Hospital who cared for our community under stress, the women who showed such bravery during pregnancy and delivery in the face of the pandemic, and the COVID-19 Unit for Research at Elmhurst for the timely research support.

## Author Contributions

Conceptualized the screening protocol, inpatient care and postpartum follow-up: BB, AD, TB, UP, RW. Conceptualized the study: SM, JM. Collected the data: RC, SM, PK. Methodology development, data analysis, interpretation: AB, SM, DM, RC, UP, PK, LN, RW, MS, BB. Contributed to the writing and editing of the manuscript: SM, UP, RC, AB, TB, AD, MS, LN, RW, KB, CW, DM, BB, RV. All authors meet ICMJE criteria for authorship and all authors read and approved the final manuscript.

## Notes

### Competing Interest Statement

The authors have declared no competing interest.

### Funding Statement

No external funding was received for this work.

